# Pilot study demonstrating changes in DNA hydroxymethylation enable detection of multiple cancers in plasma cell-free DNA

**DOI:** 10.1101/2020.01.22.20018382

**Authors:** Anna Bergamaschi, Yuhong Ning, Chin-Jen Ku, Chris Ellison, Francois Collin, Gulfem Guler, Tierney Phillips, Erin McCarthy, Wendy Wang, Michael Antoine, Aaron Scott, Paul Lloyd, Alan Ashworth, Steve Quake, Samuel Levy

## Abstract

Our study employed the detection of 5-hydroxymethyl cytosine (5hmC) profiles on cell free DNA (cfDNA) from the plasma of cancer patients using a novel enrichment technology coupled with sequencing and machine learning based classification method. These classification methods were develo^i^ped to detect the presence of disease in the plasma of cancer and control subjects. Cancer and control patient cfDNA cohorts were accrued from multiple sites consisting of 48 breast, 55 lung, 32 prostate and 53 pancreatic cancer subjects. In addition, a control cohort of 180 subjects (non-cancer) was employed to match cancer patient demographics (age, sex and smoking status) in a case-control study design.

Logistic regression methods applied to each cancer case cohort individually, with a balancing non-cancer cohort, were able to classify cancer and control samples with measurably high performance. Measures of predictive performance by using 5-fold cross validation coupled with out-of-fold area under the curve (AUC) measures were established for breast, lung, pancreatic and prostate cancer to be 0.89, 0.84, 0.95 and 0.83 respectively. The genes defining each of these predictive models were enriched for pathways relevant to disease specific etiology, notably in the control of gene regulation in these same pathways. The breast cancer cohort consisted primarily of stage I and II patients, including tumors < 2 cm and these samples exhibited a high cancer probability score. This suggests that the 5hmC derived classification methodology may yield epigenomic detection of early stage disease in plasma. Same observation was made for the pancreatic dataset where >50% of cancers were stage I and II and showed the highest cancer probability score.

## INTRODUCTION

Detection of point mutations or copy number variations in cfDNA or circulating tumor DNA (ctDNA) has revolutionized the molecular characterization of tumors. However, strategies which interrogate dynamic and temporal genome-wide signals in a liquid biopsy context may address precision medicine’s needs to improve cancer care ^1^. Exploiting the epigenetic regulation of gene expression exemplifies one systems biology approach to deciphering cancer pathways for biomedical applications. Genome-wide mapping of methylation or hydroxymethylation of cytosine residues (5mC and 5hmC) in a CpG context from tissues and cfDNA has clarified the regulatory effects of 5mC or 5hmC. These suppressive or activating effects, for 5mC and 5hmC respectively, on gene expression in cells of diverse tumor types and their inflamed microenvironments ^2–4^, have yielded new therapeutic targets^5,6^ and candidate biomarkers potentially useful for clinical decision-making^7–10^. Such investigations of non-small cell lung cancer (NSCLC) have clarified the contribution of 5mC and 5hmC at gene loci and promoter regions or CpG islands to tumor heterogeneity^11^, pre-malignant lesions^12^ and regulation of PDL1 expression^13^. Statistical analysis and machine learning approaches incorporating methylation states have yielded candidate algorithms for discrimination of clinical cancer states^8,14^. Studies of breast malignancies^15–17^ and pancreatic cancer have identified similar perturbations of 5mC or 5hmC in tissue or cfDNA. Comprehensive analyses of multiple cancer types have also yielded a candidate pan-cancer plasma cfDNA methylation panel associated with clinical outcomes^20-21^.

Development of cfDNA 5hmC biomarkers for cancer management must also address the molecular complexity of individual patients since each comprises a blend of demographic and/or clinical conditions known to impact epigenetic methylation states. Specifically, patients with cancer reflect a population segment associated with different age-dependent risk profiles with a high prevalence of both obesity and tobacco use. Analyses of peripheral blood mononuclear cells (PBMCs)^22^ and whole blood cells^23^ in European and Canadian populations, respectively, have demonstrated an age-associated decline of 5hmC levels. In addition, the metabolic, physiologic and inflammatory effects of obesity, defined as a body-mass index (BMI) of 30 or higher^24^, are associated with BMI-related changes in whole blood genomic DNA methylation levels at 187 gene loci^25^ and CpG loci-specific methylation related to BMI and waist circumference metrics^26^. Cigarette smoking has well-established associations with the development of diverse diseases and multiple cancer types^27^, including a role for DNA methylation in oncogenic pathways influenced by tobacco^28–30^. In addition to the identification of candidate hypomethylated CpG sites which may mediate the effects of tobacco use on lung cancer^31^, cigarette smoking also impacts genome-wide methylation broadly that persists at a variety of loci for many years after smoking cessation^32^. Importantly, the discrepancies in the composition and discriminatory performance of cfDNA 5hmC molecular classifiers for NSCLC (8, 14) illustrate the need to consider not only tumor histologic subtypes and pathologic staging, but also cohort-specific demographic metrics and disease co-morbidities in the interpretation of genome-wide methylation data. Whether the epigenetic impact identified previously for age, obesity and smoking might also impact cfDNA cytosine methylation states, as well as the relationship between cfDNA 5mc and 5hmC levels^33^ remain unknown. However, these may be collectively important challenges to overcome in the development of biomedical applications of 5hmC classifiers.

In our prior work, we identified enrichment of cfDNA 5hmC signatures in exons, 3’UTRs and transcription termination sites in a disease- and stage-specific manner in a case-control study of pancreatic ductal adenocarcinoma. To further investigate the clinical complexity of cancer epigenetic mechanisms mediated by the hydroxymethylome, we performed genome-wide interrogation of plasma cfDNA 5hmC levels to identify candidate hydroxymethlated genes or genomics regions. We focused our study in three cancer types, notably, (i) in women with infiltrating ductal carcinoma (IDC), (ii) in lung cancer patients across the spectrum of pathologic stage, and (iii) in prostate and pancreatic adenocarcinoma patients early stage using a case-control study design.

To control for potential confounding effects of clinical states, study cohorts were selectively matched for age, obesity (using established BMI metrics), and tobacco exposure using smoking status (current, former, never) and pack-years (defined as the product of the number of packs of cigarettes smoked daily by the number of years smoked). A machine learning framework was used to train and validate prediction models to enable both the binary and multiple classification of different cancer states. The data demonstrate that multi-gene or multi-loci 5hmC signatures appear to be specific for each cancer, i.e., organ and subject cohorts, and further that discrete classifiers can be found to distinguish amongst the three tumor types. This suggests that large scale studies of 5hmC cfDNA levels in cancer subjects using clinically well-annotated plasma specimens may enable the derivation of molecular signatures to classify and/or stratify patient cohorts along the management spectrum from early detection, i.e., screening, and early diagnosis to predictions of therapeutic responsiveness and disease recurrence.

## Material and Methods

### Clinical cohorts and study design

A case-control study was performed using plasma obtained from subjects without (termed control) and with cancer patients diagnosed with breast, lung, pancreas and prostate who provided informed consent and contributed biospecimens in studies approved by the Institutional Review Boards (IRBs) at participating sites in the United States. Plasma samples for the control cohort were obtained from subjects enrolled prospectively at up to 20 sites in the United States, following review and approval of the study protocol by each site’s participating investigator(s).

### Cancer cohort

Plasma samples for the cancer cohort were obtained from subjects who had undergone management of breast, colon or lung in the United States, and also provided consent for use of blood specimens for archival storage and retrospective analyses. Criteria for subject eligibility for inclusion in the analysis included age greater than or equal to 21 years for all subjects, with additional requirements for the cancer cohort including: 1) no cancer treatment, e.g., surgical, chemotherapy, immunotherapy, targeted therapy, or radiation therapy, prior to study enrollment and blood specimen acquisition; and 2) a confirmed pathologic diagnosis.

### Control cohort

Subject exclusion criteria for the non-cancer cohort also included any of the following: prior cancer diagnosis within prior six months; surgery or invasive procedure requiring general anesthesia within prior month; non-cancer systemic therapy associated with molecularly targeted immune modulation; concurrent or prior pregnancy within previous 12 months; history of organ tissue transplantation; history of blood product transfusion within one month; and major trauma within six months. Clinical data required for all subjects included age, gender, smoking history, and both tissue pathology and grade, and were managed in accordance with the guidance established by the Health Insurance Portability and Accountability Act (HIPAA) of 1996 to ensure subject privacy.

### Plasma collection

Plasma was isolated from whole blood specimens obtained by routine venous phlebotomy at the time of subject enrollment. For both cancer and control subjects, whole blood was collected in Cell-Free DNA BCT^®^ tubes according to the manufacturer’s protocol (Streck, La Vista, NE) (https://www.streck.com/collection/cell-free-dna-bct/). Tubes were maintained at 15 °C to 25 °C with plasma separation performed within 24 h of phlebotomy by centrifugation of whole blood at 1600 × g for 10 min at RT, followed by transfer of the plasma layer to a new tube for centrifugation at 3500 × g for 10 min. Plasma was aliquoted for subsequent cfDNA isolation or storage at −80°C.

### cfDNA isolation

cfDNA was isolated using the QIAamp Circulating Nucleic Acid Kit (QIAGEN, Germantown, MD) following the manufacturer’s protocol excepting the omission of carrier RNA during cfDNA extraction. Four milliliter plasma volumes were used for cfDNA extraction. Eluates were collected in a volume of 60 µl buffer. All cfDNA extracts were quantified by Qubit dsDNA High Sensitivity Assay (Thermo Fisher Scientific, Waltham, MA) and quality of fragment sizes by Tape Station (Agilent Technologies Inc, Santa Clara, CA).

### 5-hydroxymethyl Cytosine (5hmC) assay enrichment

Sequencing library preparation and 5hmC enrichment was performed as described previously^8^. cfDNA was normalized to 10 ng total input for each assay and ligated to sequencing adapters. The adapter ligated library was partitioned 80:20 to enable 5hmC enrichment and whole genome sequencing to be performed on each partition. 5hmC bases were biotinylated via a two-step chemistry and subsequently enriched by binding to Dynabeads M270 Streptavidin (Thermo Fisher Scientific, Waltham, MA). All libraries were quantified by Bioanalyzer dsDNA High Sensitivity assay (Agilent Technologies Inc, Santa Clara, CA) and Qubit dsDNA High Sensitivity Assay (Thermo Fisher Scientific, Waltham, MA) and normalized in preparation for sequencing.

### DNA sequencing and alignment

DNA sequencing was performed according to manufacturer’s recommendations with 75 base-pair, paired-end sequencing using a NextSeq550 instrument with version 2 reagent chemistry (Illumina, San Diego, CA). Twenty-four libraries were sequenced per flowcell,to yield approach 20 millon paired end reads, and raw data processing and demultiplexing was performed using the Illumina BaseSpace Sequence Hub to generate sample-specific FASTQ output. Sequencing reads were aligned to the hg19 reference genome using BWA-MEM with default parameters.

### 5hmC Profile Quality Control

Subsequent to alignment, we performed a variety of quality control checks on the 5hmC profiles. Additionally, the 5hmC variability for each gene, in each sample, was compared to the median count for the same genes in the control population. Each sample’s range of 5hmC count was thus expressed as a relative log representation (RLR) to the control cohort median 5hmC values.

### Predictive Modelling

For the purpose of assessing the feasibility of building classifiers that can discriminate between cancer type and or all cancers and non-cancer samples based on the 5hmC representation of gene bodies, we evaluated the performance of two forms of regularized logistic regression models, namely the Lasso and the Elastic net, which are commonly used in the classification context, where the number of examples are few and the number of features are large. See Friedman et al.^36^ for a description of the general Elastic net procedure. Software implementation of these methods can be found at https://cran.r-project.org/web/packages/glmnet/index.html. To remove weakly represented genes, we excluded genes that did not have greater than 3 counts per million reads in at least 20 samples. This filter excludes roughly 12% of the genes. Before any fitting, genes were filtered to include the 65% of the most variable genes for model fitting task.

We employed lasso and elastic net, which control the level of regularization used in the fit by requiring to specify hyper-parameters. Hyper-parameters were selected based on out-of-fold performance on 30 repetitions of 10-fold cross-validated analysis of the training data. Out-of-fold assessments are based on the samples in the left-out fold at each step of the cross-validated analysis. The same approach was employed when including PCAWG ^37^ enhancers in the model.

## RESULTS

### Clinical cohort and Study Design

Plasma specimens were collected from 180 presumably healthy controls and 48 breast, 55 lung, 53 pancreatic and 32 prostate cancer patients. The schematic summary of cancer cohorts, with the corresponding matched control cohorts, can be found in Fig 1. Study inclusion criteria for the cancer and controls are reported in the Material and Methods section and cohorts clinico-pathological features description are listed in Table 1.

**Table 1.** Cohorts clinico-pathological features. Statistical analysis of differentially represented genes and prediction models in the lung cancer cohort were corrected for smoking status differences. All cohorts were age matched.

|  | Breast Dataset |  | Lung Dataset |  | Pancreas Dataset |  | Prostate Dataset |  |
| --- | --- | --- | --- | --- | --- | --- | --- | --- |
|  | Cancer | Control | Cancer | Control | Cancer | Control | Cancer | Control |
| Total, n | 48 | 40 | 55 | 55 | 53 | 53 | 32 | 32 |
| Age | 57.4 | 58 | 70.4 | 69.9 | 66.4 | 66.4 | 70.3 | 70.6 |
| Sex (Female) | 48 | 40 | 31 | 24 | 30 | 24 | 0 | 0 |
| Stage |  |  |  |  |  |  |  |  |
| NA | 1 |  | 5 |  | 2 |  |  |  |
| I | 25 |  | 9 |  | 11 |  | 18 |  |
| II | 17 |  | 1 |  | 19 |  | 12 |  |
| III | 4 |  | 23 |  | 6 |  | 1 |  |
| IV | 1 |  | 17 |  | 15 |  | 1 |  |
| Histology |  |  |  |  |  |  |  |  |
| Invasive Adenocarcinoma | 48 |  | 35 |  | 53 |  | 32 |  |
| Squamous Cell Carcinoma | - |  | 20 |  |  |  |  |  |
| Smoking |  |  |  |  |  |  |  |  |
| Current | 2 (4.2%) | 4 (10%) | 15 (27.2%) | 15 (27.2%) | 5 (25%) | 4(10%) | 3 (9.4%) | 4 (12.5%) |
| Former | 8 (16.7) | 6 (15%) | 32 (58.2%) | 32 (58.2%) | 3 (15%) | 15(38%) | 9 (28.1%) | 14 (43.75) |
| Never | 38 (79.1%) | 30 (75%) | 8 (14.6%) | 8 (14.6%) | 12 (60%) | 21 (52%) | 20 (62.5%) | 14 (43.75) |
| BMI |  |  |  |  |  |  |  |  |
| median | 30.7 | 29.5 | 31.1 | 28.4 | 25.4 | 28.6 | 27.8 | 28.7 |

**Fig 1.**
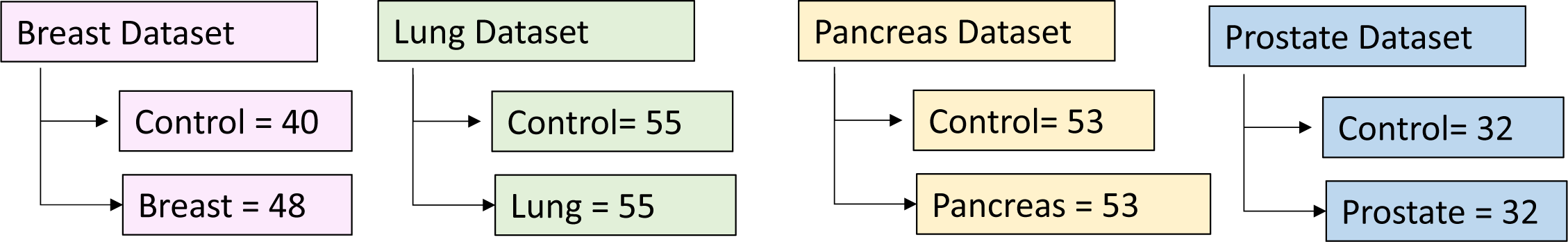
Summary of Cohorts included in the study. Schematic depicting study datasets included in the study for breast, lung, pancreas and prostate datasets including number of controls used. Case-control design employed 1:1 ratio (control:cases)

### Cancer specific predictive models for the detection of breast, lung and pancreatic cancer in cfDNA

We performed regularized logistic regression analysis in order to determine whether gene-based features are present in the breast, lung, pancreas and prostate and non-cancer cohorts to enable the classification of patient samples.

For all datasets we employed 3 type of analysis: i) 65% of the genes with the most variable 5hmC density for model selection only; ii) PCAWG enhancers only and iii) a combined gene and enhancers analysis. As a regularization method we use Elastic net which require specifying hyper-parameters for controlling the level of regularization used in the fit. These hyper-parameters were selected based on out-of-fold performance on 30 repetitions of 5-fold cross-validated analysis of the training data. Out-of-fold assessments are based on the samples in the left-out fold at each step of the cross-validated analysis. Performance of all three analysis performances can be found in Table 2.

**Table 2.** Area Under the Curve (auROC) for all cancer types

|  | Gene Body<br>Only | Enhancers<br>Only | Gene body +<br>Enhancers |
| --- | --- | --- | --- |
| Breast | 0.89 | 0.72 | 0.83 |
| Lung | 0.79 | 0.78 | 0.84 |
| Pancreas | 0.93 | 0.92 | 0.95 |
| Prostate | 0.83 | 0.83 | 0.72 |

The training set for genes only yielded an out-of-fold performance metric, Area Under Curve (AUC) of 0.89, 0.79, 0.93 and 0.83 respectively for breast, lung, pancreatic and prostate respectively.

The training set for enhancers only yielded an out-of-fold performance metric AUC of 0.72, 0.8, 0.92 and 0.82 respectively for breast, lung, pancreatic and prostate respectively.

The training set for genes and enhancers yielded an out-of-fold performance metric, AUC of 0.83, 0.86, 0.95 and 0.73 respectively for breast, lung, pancreatic and prostate respectively. Amon the 3 different methods the best performance for breast was found with using gene-only analysis AUC= 0.89 with an interquartile range (AUC-IQR) of the 30 five-fold cross validation runs were, for breast, 0.72 – 0.92 (Fig 2A).

**Fig 2.**
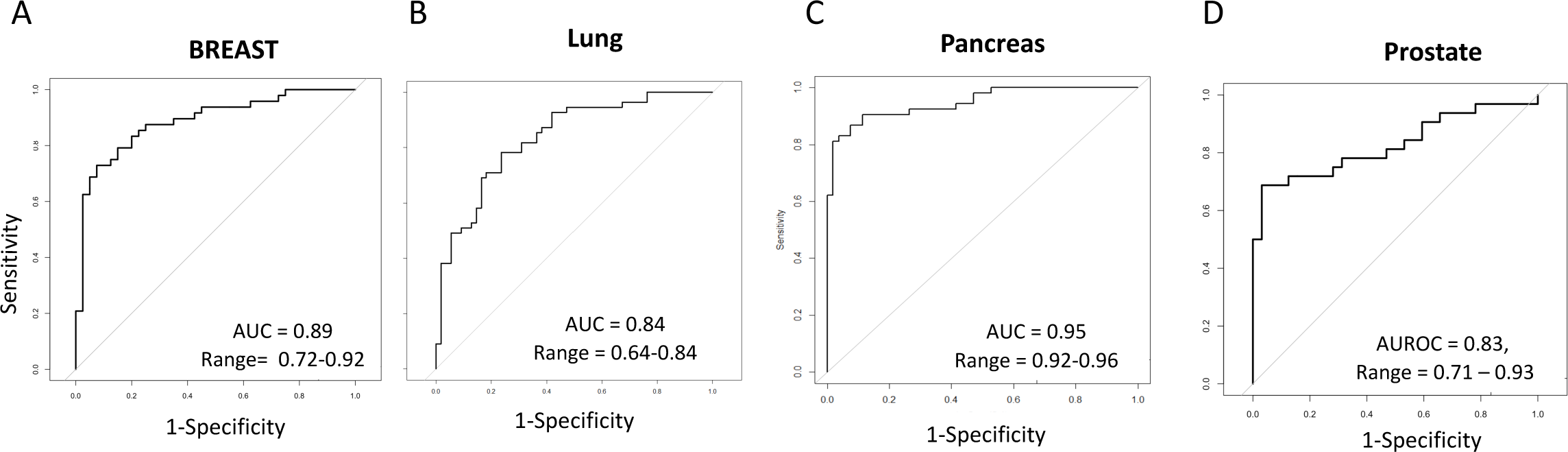
Identification of a 5hmC signature that differentiates breast, lung, pancreas and prostate cancers from controls. Logistic regression algorithms were trained for each cohort. Correction for smoking was performed in the lung cohort.

The best performance for lung and pancreas was found using genes plus enhancers AUC respectively of 0.84 and 0.95. AUC-IQR for lung and pancreas were 0.64-0.84, pancreas 0.92-0.96.

Performace for prostate cancer was the highest when genes-only and enhancers-only analysis was employed (AUC 0.83 with and AUC-IQR of 0.71 – 0.93).

Summary of sensitivity performance is found in Table 3. Sensitivity, at 95% specificity, was calculated for each cancers for the highest model. Pancreas prediciton model had the highest sensitivity (83%) followed by beast (69%), prostate (64%) and lung (49%).

**Table 3.** Cancer cohorts sensitivity calculated at 95% specificity

| Cancer type | 95% Specificity |
| --- | --- |
| Breast | 69% |
| Lung | 49% |
| Pancreas | 83% |
| Prostate | 64% |

### Cancer predictive models are associated with enrichment of specific biological pathways

We sought to identify whether the features identified in the 4 cancer prediction models were associated with any relevant cancer biology.

First, we noted that in breast cancer, the majority of early stage with small size tumors (<2cm) had the highest cancer predicted probability (Fig 3a). Same observation was made for the pancreas dataset (Fig. D), while for prostate and lung the highest prediction scores are spread across diffent stages (Fig. 3C-E).

**Fig 3.**
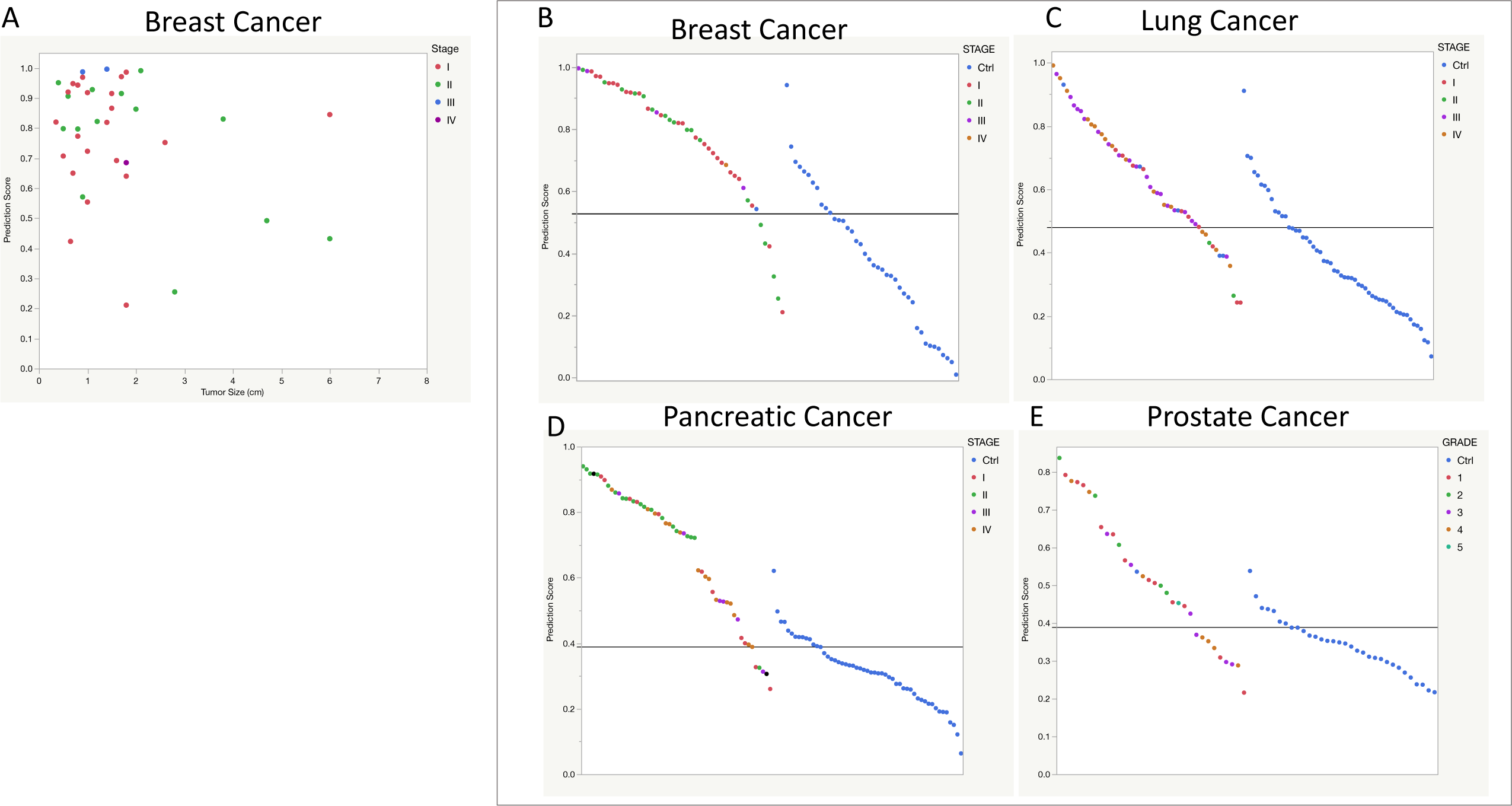
Distribution of classification scores for individual cancer and control cohorts depicting. A threshold was set by calculating the third quartile of control samples. Cancer samples are color-coded based on stage.

Next, we inspected genes defined by the predictive models to determine if any biologically relevant pathways specific for breast, lung, pancreas and prostate cancer or tumorigenesis more generally, was enriched using mSigDB^37^ and KEGG pathway analysis. 5hmC predictive genes in breast were enriched in BRCA1 network, hypoxia genes signaling and early estrogen response genes (Table 4).

**Table 4.** Enriched genesets in the breast cancer prediction model

| Description | P-value |
| --- | --- |
| HALLMARK_ESTROGEN_RESPONSE_EARLY | 0.0001 |
| BRCA1_PCC_NETWORK | 0.01 |
| HALLMARK_HYPOXIA | 0.04 |

Using KEGG pathway analysis, predictive model in lung cancer, was found to be enriched with response to necrosis factor and EGFR signaling pathway genes (Fig. 4A). Of Interest, KEGG pathway analysis indeitified in the pancreatic cancer model Notch signaling pathway to be significantly enriched (Fig. 4B).

**Fig 4.**
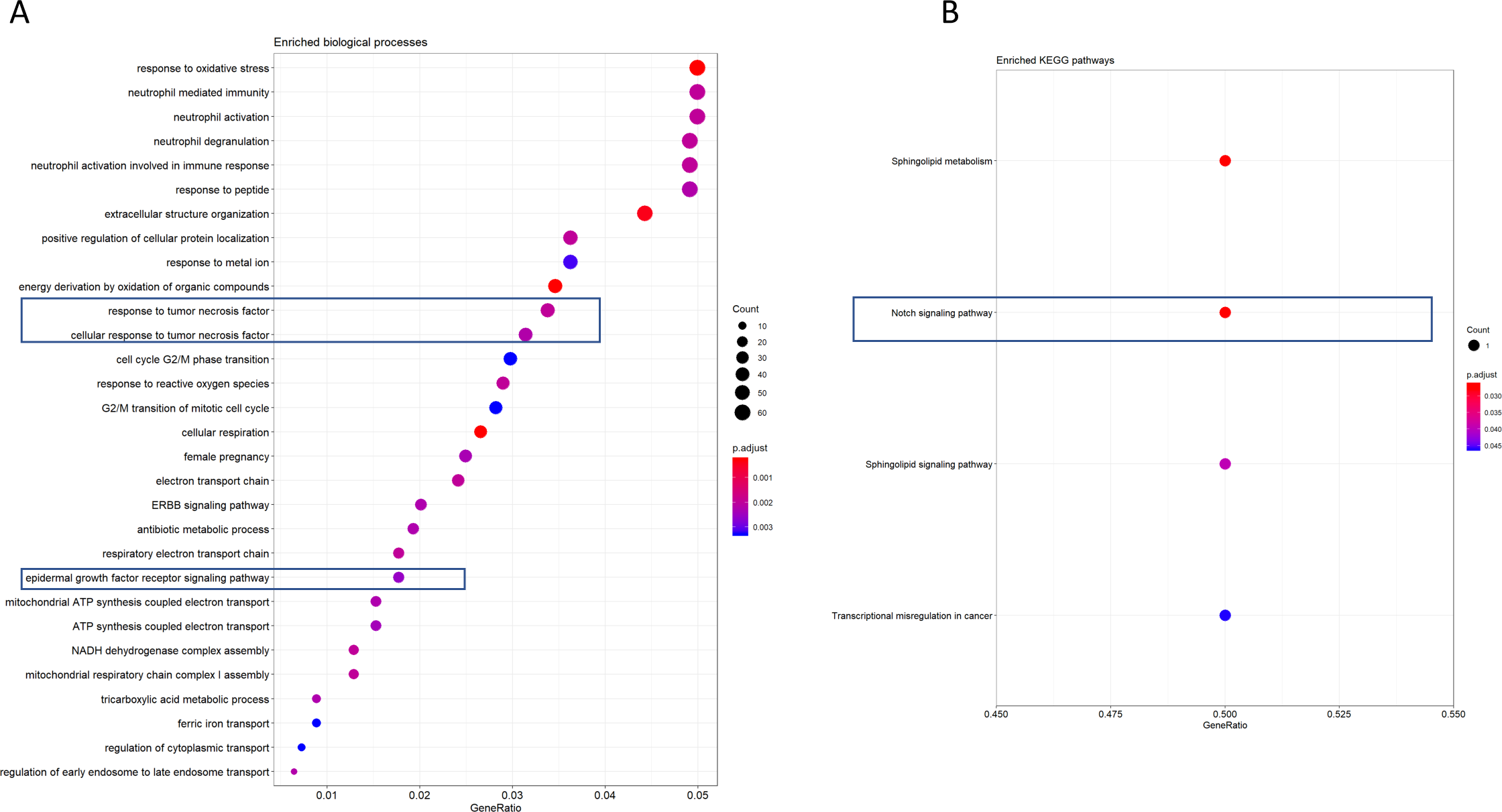
Identification of KEGG pathways identified by the A)lung cancer B) Pancreatic prediction model.

## DISCUSSION

The aim of this pilot study was to evaluate whether cfDNA specific hydroxymethyl cytosine-based profiles can be employed to discover candidate biomarkers that are specific for the detection of breast, lung, pancreas and prostate cancer. This approach identified predictive hydroxymethylated genes and gene loci and, in breast, lung, pancreas and prostate cancer patients. These predictive genes were found involved in known cancer pathways such as BRCA, and hypoxia in breast cancer, EGFR and TNF and Pholipase A2 in pancreatic cancer. In breast cancer, it was possible to discover a set of genes that contributed to predictive models with good performance in separating cancer from controls, AUC = 0.89. High probability scores from the gene-based prediction model in breast cancer are associated with tumor size < 2 cm, suggesting that despite the apparent absence of mutation in the cfDNA, that aberrant biology and clinical characteristics can be identified through the biomarker signatures (Fig 3A). This lends support to the hypothesis that relevant underlying biological transformations of early stage oncological disease are being employed in cancer discrimination.

Previous studies in breast cancer have shown that a global decrease in 5hmC occurs and is associated with a poor prognosis^16,38^. Our data show that 5hmC changes in the cfDNA from plasma has more nuanced changes, including increases and decreases in 5hmC, that bear the molecular hallmarks of breast cancer.

Several cancer specific pathways were found to be activated in the breast cancer predictive model, by employing the contributing genes in the model, to measure pathway enrichment directly. Of note, amongst the enrichment was the hypoxia pathways. Changes in the cellular environment through hypoxia have been identified in breast cancer^36^ and associated with poor prognosis. Hypoxia has been shown to control methylation through TET1/3 deregulation^39^ which would result in altered 5hmC patterns. Through the detection of these modulated 5hmC profiles in our breast cancer cohort, as found in the predictive model, we have been able to show that genes involved in the mediation hypoxia are implicated.

The best lung cancer classification, with an AUC = 0.84, was identified by genes and enhancers, possibly indicating that the samples in the datasets may have a high transcriptional activity compared to control samples.

Futher, our lung results compared favorably to the data of Zhang et al^14^, despite the fact that different training methods were employed as well as the apparent absence of out-of-fold validation in the latter study. Another source of difference with Zhang et al is the lung cancer histology which may be a combination of adenocarcinoma, squamous and adenosquamous. Our study employed primarily adenocarcinoma samples (>75% lung cancer cohort).

Epidermal growth factor receptor signalling pathway, a known lung-associated pathways was found enriched in predictive genes and enhancers in the lung dataset. Its activation is a key driver of lung cancer progression and the most important target for lung cancer treatment. We also investigate pathway enriched in the pancreatic cancer and interestingly found NOTCH siganlin genes significantly enriched in the pancreatic prediction model. Indeed Notch signaling pathway has been showed to play critical roles in the development and progression of pancreatic cancer^41^.

In summary, we show here a platform technology employing 5hmC signal detection coupled with machine learning approaches to develop methodologies that can extend toward the detection of specific cancers or to distinguish amongst various cancer types. These complex problems can be tackled in a liquid biopsy context and provide the opportunity for the development of future assays of clinical utility in the earlier diagnosis of disease.

## Data Availability

Data will be made available upon acceptance of this manuscript

## REFERENCES

1. Corcoran, R. B. & Chabner, B. A. Application of Cell-free DNA Analysis to Cancer Treatment. New England Journal of Medicine 379, 1754–1765 (2018).

2. Cavalcante, R. G., Patil, S., Park, Y., Rozek, L. S. & Sartor, M. A. Integrating DNA Methylation and Hydroxymethylation Data with the Mint Pipeline. Cancer research 77, e27–e30 (2017).

3. de la Cruz, F. F. & Corcoran, R. Methylation in cell-free DNA for early cancer detection. Ann Oncol (2018). doi:10.1093/annonc/mdy134

4. Koch, A. et al. Analysis of DNA methylation in cancer: location revisited. Nat Rev Clin Oncol 15, 459–466 (2018).

5. Mohammad, H. P., Barbash, O. & Creasy, C. L. Targeting epigenetic modifications in cancer therapy: erasing the roadmap to cancer. Nat Med 25, 403–418 (2019).

6. Cho, J. et al. Epigenetic Therapeutics and Their Impact in Immunotherapy of Lung Cancer. Curr Pharmacol Reports 3, 360–373 (2017).

7. Li, W. et al. 5-Hydroxymethylcytosine signatures in circulating cell-free DNA as diagnostic biomarkers for human cancers. Cell Research 1–15 doi:10.1038/cr.2017.121

8. Song, C.-X. et al. 5-Hydroxymethylcytosine signatures in cell-free DNA provide information about tumor types and stages. Cell Res 27, 1231 (2017).

9. Pfeifer, G. P. & Szabó, P. E. Gene body profiles of 5-hydroxymethylcytosine: potential origin, function and use as a cancer biomarker. Epigenomics (2018). doi:10.2217/epi-2018-0066

10. Pfeifer, G. P., Xiong, W., Hahn, M. A. & Jin, S. The role of 5-hydroxymethylcytosine in human cancer. Cell and Tissue Research 356, 631–641 (2014).

11. Vargas, A. J. & Harris, C. C. Biomarker development in the precision medicine era: lung cancer as a case study. Nat Rev Cancer 16, nrc.2016.56 (2016).

12. Teixeira, V. H. et al. Deciphering the genomic, epigenomic, and transcriptomic landscapes of pre-invasive lung cancer lesions. Nat Med 25, 517–525 (2019).

13. Kowanetz, M. et al. Differential regulation of PD-L1 expression by immune and tumor cells in NSCLC and the response to treatment with atezolizumab (anti–PD-L1). Proc National Acad Sci 115, 201802166 (2018).

14. Zhang, J. et al. 5-Hydroxymethylome in Circulating Cell-free DNA as A Potential Biomarker for Non-small-cell Lung Cancer. Genomics, proteomics & bioinformatics 16, 187–199 (2018).

15. Wilkins, O. M. et al. Genome-wide abundance of 5-hydroxymethylcytosine in breast tissue reveals unique function in dynamic gene regulation and carcinogenesis. Biorxiv 339069 (2018). doi:10.1101/339069

16. Tsai, K.-W. et al. Reduction of global 5-hydroxymethylcytosine is a poor prognostic factor in breast cancer patients, especially for an ER/PR-negative subtype. Breast Cancer Res Tr 153, 219–234 (2015).

17. Connolly, R. M. et al. Tumor and serum DNA methylation in women receiving preoperative chemotherapy with or without vorinostat in TBCRC008. Breast Cancer Res Tr 167, 107–116 (2018).

18. Gilat, N. et al. Single-molecule quantification of 5-hydroxymethylcytosine for diagnosis of blood and colon cancers. 1–8 (2017). doi:10.1186/s13148-017-0368-9

19. Uribe-Lewis, S. et al. 5-hydroxymethylcytosine marks promoters in colon that resist DNA hypermethylation in cancer. Genome Biol 16, 69 (2015).

20. Liu, L. et al. Targeted Methylation Sequencing of Plasma Cell-free DNA for Cancer Detection and Classification. Ann Oncol Official J European Soc Medical Oncol (2018). doi:10.1093/annonc/mdy119

21. Valentini, E. et al. Analysis of the machinery and intermediates of the 5hmC-mediated DNA demethylation pathway in aging on samples from the MARK-AGE Study. Aging 8, 1896–1922 (2016).

22. Buscarlet, M., Tessier, A., Provost, S., Mollica, L. & Busque, L. Human blood cell levels of 5-hydroxymethylcytosine (5hmC) decline with age, partly related to acquired mutations in TET2. Experimental Hematology 44, 1072–1084 (2016).

23. Heymsfield, S. B. & Wadden, A. Mechanisms, Pathophysiology, and Management of Obesity. New Engl J Medicine 376, 1490–1492 (2017).

24. Wahl, S. et al. Epigenome-wide association study of body mass index, and the adverse outcomes of adiposity. Nature 541, 81 (2017).

25. Sayols-Baixeras, S. et al. DNA methylation and obesity traits: An epigenome-wide association study. The REGICOR study. Epigenetics 0–0 (2017). doi:10.1080/15592294.2017.1363951

26. Carter, B. D. et al. Smoking and Mortality — Beyond Established Causes. New Engl J Medicine 372, 631–640 (2015).

27. Lee, K. & Pausova, Z. Cigarette smoking and DNA methylation. Front Genet 4, 132 (2013).

28. Zhang, Y. et al. DNA methylation signatures in peripheral blood strongly predict all-cause mortality. Nat Commun 8, 14617 (2017).

29. Ma, Y. & Li, M. D. Establishment of a Strong Link Between Smoking and Cancer Pathogenesis through DNA Methylation Analysis. Sci Rep-uk 7, 1811 (2017).

30. Fasanelli, F. et al. Hypomethylation of smoking-related genes is associated with future lung cancer in four prospective cohorts. Nat Commun 6, 10192 (2015).

31. Joehanes, R. et al. Epigenetic Signatures of Cigarette Smoking. Circulation Cardiovasc Genetics 9, 436–447 (2018).

32. Li, L., Gao, Y., Wu, Q., Cheng, A. & Yip, K. Y. New guidelines for DNA methylome studies regarding 5-hydroxymethylcytosine for understanding transcriptional regulation. Genome Res 29, 543–553 (2019).

33. Law, C. W., Alhamdoosh, M., Su, S., Smyth, G. K. & Ritchie, M. E. RNA-seq analysis is easy as 1-2-3 with limma, Glimma and edgeR. F1000Research 5, 1408 (2016).

34. Benjamini, Y. & Hochberg, Y. Controlling the false discovery rate: a practical and powerful approach to multiple testing. Journal of the royal statistical society. Series B (Methodological) 289–300 (1995).

35. Semenza, G. L. Molecular mechanisms mediating metastasis of hypoxic breast cancer cells. Trends Mol Med 18, 534–543 (2012).

36. Chang, K., Creighton, C., Davis, C. et al. The Cancer Genome Atlas Pan-Cancer analysis project. Nat Genet 45, 1113–1120 (2013) doi:10.1038/ng.2764

37. Subramanian, A. et al. Gene set enrichment analysis: A knowledge-based approach for interpreting genome-wide expression profiles. Proceedings of the National Academy of Sciences of the United States of America 102, 15545–15550 (2005).

38. Sant, D. W. et al. Vitamin C promotes apoptosis in breast cancer cells by increasing TRAIL expression. Scientific Reports 8, 5306 (2018).

39. Wu, M.-Z. et al. Hypoxia Drives Breast Tumor Malignancy through a TET–TNFα–p38–MAPK Signaling Axis. Cancer Res 75, 3912–3924 (2015).

40. Cohen, J. D. et al. Detection and localization of surgically resectable cancers with a multi-analyte blood test. Science 3247, eaar3247

41. Gao J, Long B, Wang Z. Role of Notch signaling pathway in pancreatic cancer. Am J Cancer Res. 2017;7(2):173–186.

